# Association of rare *APOE* missense variants with Alzheimer’s disease in the Japanese population

**DOI:** 10.1101/2024.11.05.24316515

**Authors:** Akinori Miyashita, Ai Obinata, Norikazu Hara, Risa Mitsumori, Daita Kaneda, Yoshio Hashizume, Terunori Sano, Masaki Takao, Ramil Gabdulkhaev, Mari Tada, Akiyoshi Kakita, Akira Arakawa, Maho Morishima, Shigeo Murayama, Yuko Saito, Hiroyuki Hatsuta, Tomoyasu Matsubara, Akio Akagi, Yuichi Riku, Hiroaki Miyahara, Jun Sone, Mari Yoshida, Haruyasu Yamaguchi, Tamao Tsukie, Mai Hasegawa, Kensaku Kasuga, Masataka Kikuchi, Ryozo Kuwano, Takeshi Iwatsubo, Japanese Alzheimer’s Disease Neuroimaging Initiative, Shumpei Niida, Kouichi Ozaki, Takeshi Ikeuchi

## Abstract

**Background:** *APOE* is a major susceptibility gene for Alzheimer’s disease (AD). Recent studies in Europe and the US have identified rare missense variants (RMVs) in *APOE* which are significantly associated with AD. However, little is known regarding *APOE* RMVs in East Asians, including the Japanese, and their association with AD and lipid metabolism.

**Objective:** To identify *APOE* RMVs in the Japanese population and investigate their association with AD and lipid metabolism, including low-density lipoprotein cholesterol levels.

**Subjects:** *APOE* RMVs were explored in the NIG (2,589 subjects) and ToMMo (3,307 subjects) cohorts. A case-control study was performed involving 6,471 AD and 20,270 control subjects.

**Methods:** Sanger sequencing or whole-exome sequencing was performed on NIG subjects. We used genotype data from ToMMo. *APOE* RMV frequencies in the Japanese were compared with various ethnic populations. Associations among *APOE* RMV genotypes, AD, and lipoproteins were examined.

**Results:** Fourteen RMVs were identified (minor allele frequency 0.02 – 0.73%), 10 of which were specifically found in East Asians. Five RMVs previously reported overseas were not detected in Japanese individuals. Two RMVs (rs140808909 and rs190853081) exhibiting complete linkage disequilibrium were associated with AD protective effects: *P*_Bonferroni_ = 3.63E-02, OR (95% CI) = 0.70 (0.54–0.92). No significant differences in cholesterol levels were observed between RMV carriers and non-carriers.

**Conclusions:** *APOE* RMVs were identified in the Japanese population, with two showing potential protective effects against AD. Further studies involving larger cohorts are required to confirm our findings and investigate the role of *APOE* RMVs in AD and lipid metabolism.

## Introduction

Alzheimer’s disease (AD) is the leading cause of dementia in elderly individuals. AD is a multifactorial disease resulting from the interaction between environmental factors and individual genetic susceptibility. A large twin epidemiological study in Sweden estimated the heritability of AD to be 60–80%. ^1^ Multiple loci are suggested to contribute to the onset of AD. ^2^ This indicates that genetic factors play a significant role in the development and progression of AD. Large-scale genome-wide association studies (GWAS) have identified 90 susceptibility genes/loci for AD. ^3^ Recent studies have focused on applying polygenic risk score analysis ^4–6^ and polygenic resilience score analysis ^7^, which integrate multiple AD susceptibility variants to evaluate individual risks, with potential clinical applications being explored.

Among the AD susceptibility genes/loci identified to date, *APOE* is the most powerful and explanatory gene across various populations. ^8–11^ A search for “Alzheimer” on the GWAS Catalog website ^12^ reveals that the genomic region containing *APOE* shows a remarkably strong association and is a “multiple hit locus” with numerous variants associated with AD. Recently, rare missense variants (RMVs) of *APOE* have been reported to be associated with AD, shedding light on new links between *APOE* and AD. For example, RMVs such as p.L46P (p.L28P; rs769452 [Pittsburgh]) ^13^, p.R154S (p.R136S; rs121918393 [Christchurch]) ^14^, p.R163C (p.R145C; rs769455) ^15^, p.V254E (p.V236E; rs199768005 [Jacksonville]) ^16^, and p.R269G (p.R251G; rs267606661) ^17^ have been reported in Western populations, showing resistance to amyloid-ß (Aß), phosphorylated tau protein, or the *APOE-e4* allele. Rasmussen *et al*. ^18^ identified 20 *APOE* RMVs in an analysis involving more than 100,000 participants and demonstrated their association with plasma *APOE* levels and AD onset.

However, there have been few reports regarding the identification and significance of *APOE* RMVs in the Japanese population. Therefore, this study was the first attempt at identifying *APOE* RMVs in the Japanese population. Here, we examined the genetic association between the identified RMVs and AD. Moreover, we investigated whether there were differences in peripheral blood lipoprotein levels between carriers and non-carriers of these RMVs.

## Subjects And Methods

### Subjects for identifying APOE RMVs

We explored *APOE* RMVs in two Japanese cohorts **(Table 1)**. The first cohort, referred to as the NIG, consisted of 2,589 participants (1,150 females and 1,439 males) from medical and research institutions across Japan. This cohort was primarily established for research-level genetic diagnoses, including AD and various other neurological disorders. Of these, 826 cases were based on clinical diagnoses, and 1,763 were based on pathological diagnoses. Additionally, 508 of the 2,589 NIG subjects were derived from the Japanese Alzheimer’s Disease Neuroimaging Initiative (J-ADNI). ^19^ Sanger sequencing (SS), whole-exome sequencing (WES), and a combination of both (SS + WES) were performed on 839, 1,509, and 241 subjects, respectively. The second cohort, referred to as ToMMo, consisted of 3,307 community-dwelling healthy subjects (1,810 females and 1,497 males) recruited by the Tohoku Medical Megabank Organization. ^20^ In compliance with the data usage policy (Research ID: 2019-0074), individual genotype data for 176 variants within a genomic region of approximately 7.2 kb, including *APOE* (positioned at 45,407,254 – 45,414,449 bp in GRCh37/hg19), were obtained. The total number of subjects from both cohorts was 5,896 (2,960 females and 2,936 males), with participants aged ≥20 years (mean: 64.1 years, median: 66.0 years, range: 20–111 years).

**Table 1.**
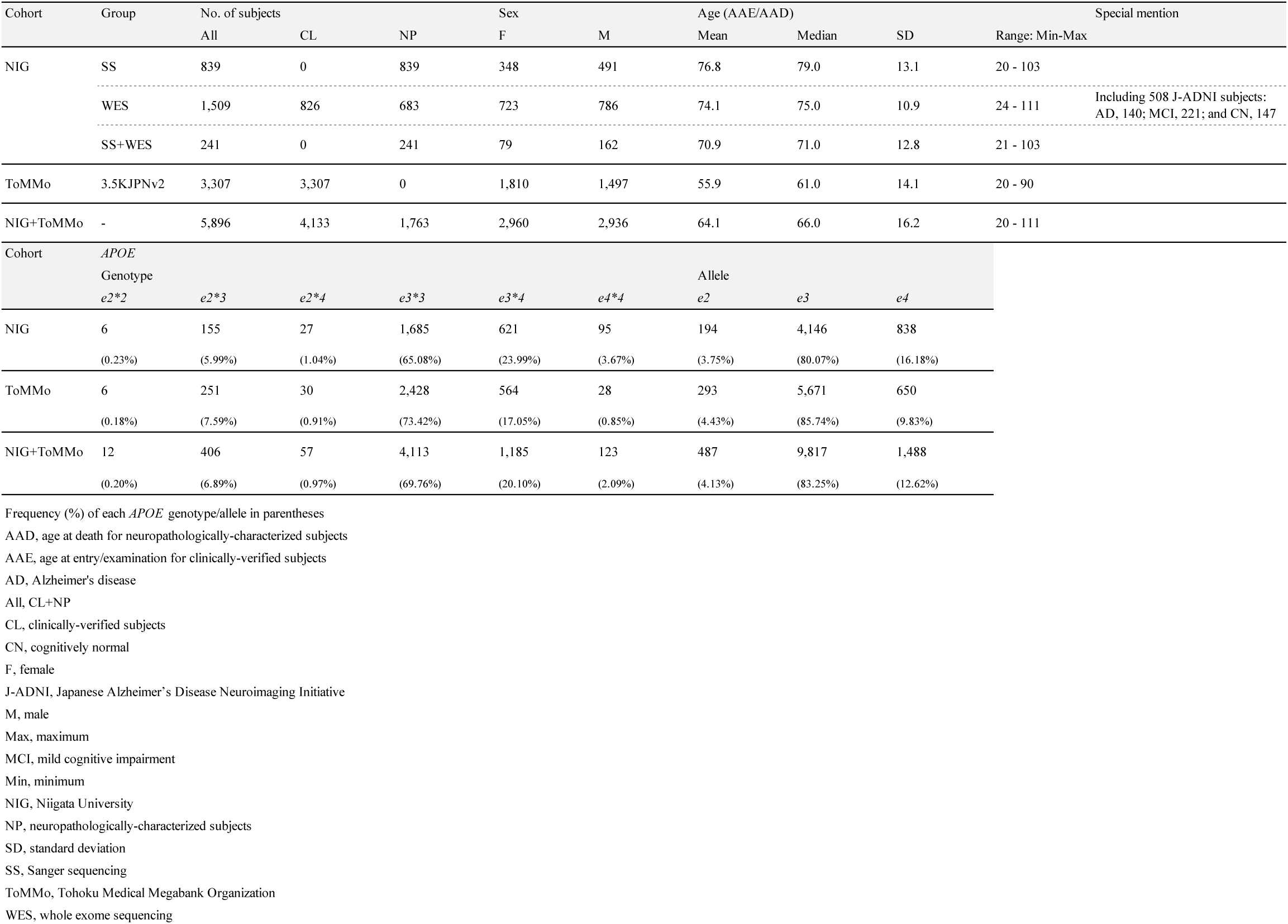
The information on subjects used in the exploration of *APOE* RMVs.

This study was approved and conducted according to the ethical guidelines of the Ethics Committee of Niigata University and all participating medical and research institutions (approval number of the Genetic Ethics Review Committee, Niigata University: G2017-0012, G2018-0034, 2018-0409, and G2022-0009).

### SS for APOE

SS was performed in 1,080 subjects from the NIG cohort (839 SS; 241 SS + WES) **(Table 1)**. Genomic DNA was extracted from peripheral blood or autopsied brain tissues and quantified using the NanoDrop spectrophotometer (Thermo Fisher Scientific). DNA quality was assessed by calculating the DNA integrity number (Agilent Technologies). The primers used for sequencing the entire coding region of *APOE* is provided in **Supplementary Table 1**.

### WES

WES was performed on 1,750 participants from the NIG (1,509 for WES and 241 for SS + WES) **(Table 1)**. Genomic DNA was used to prepare an exome library using the SureSelect Human All Exon V6 Kit (Agilent Technologies). The library was sequenced on an NovaSeq 6000 platform (Illumina) in 151-cycle paired-end mode. Sequence reads were processed with fastp (v.0.19.5) using the default settings for quality control and adapter trimming. The cleaned reads were then mapped to the human reference genome (hg38) using BWA-MEM (v.0.7.15-r1140) with default parameters. Subsequent analyses, including read processing, variant calling, and filtration, were performed following GATK4 Best Practices. The resulting variant call sets were annotated using snpEff (v.4.3t) and stored in an SQLite database using GEMINI (v.0.20.1).

### Determination of six common APOE genotypes

We determined six common *APOE* genotypes (*e2*2*, *e2*3*, *e2*4*, *e3*3*, *e3*4*, and *e4*4*) in all 5,896 subjects from the NIG and ToMMo cohorts **(Table 1)** based on nucleotide combinations (haplotypes) of the common missense variants rs429358 (T>C: p.C130R [p.C112R]) and rs7412 (C>T: p.R176C [p.R158C]) in *APOE*. For the NIG cohort, these genotypes were determined using both the SS and TaqMan methods. Information regarding the TaqMan assays used (TaqMan assay IDs: 19AyD0001 for rs429358 and 19AyD0002 for rs7412) is provided in **Supplementary Table 1**. For the ToMMo cohort, genotypes were determined based on digital data (genotype: 0/0, 0/1, and 1/1). The results are presented in **Table 1**. The risk effects of *APOE* genotypes and alleles in Japanese patients with AD are shown in **Supplementary Figure 1**. *APOE* genotype and allele information reported by Asanomi *et al*. ^21^ was used in this analysis.

### Case-control study

We conducted a case-control study to examine whether the identified *APOE* RMVs were genetically associated with AD. Information concerning the subjects used in this analysis is provided in **Table 3**, including sex, age (age at testing/death), and *APOE* genotype. Genomic variants for each subject were comprehensively genotyped using an Asian Screening Array (ASA) (Illumina). The number of samples analyzed by the ASA was 6,471 patients with AD and 20,270 control subjects.

**Table 2.**
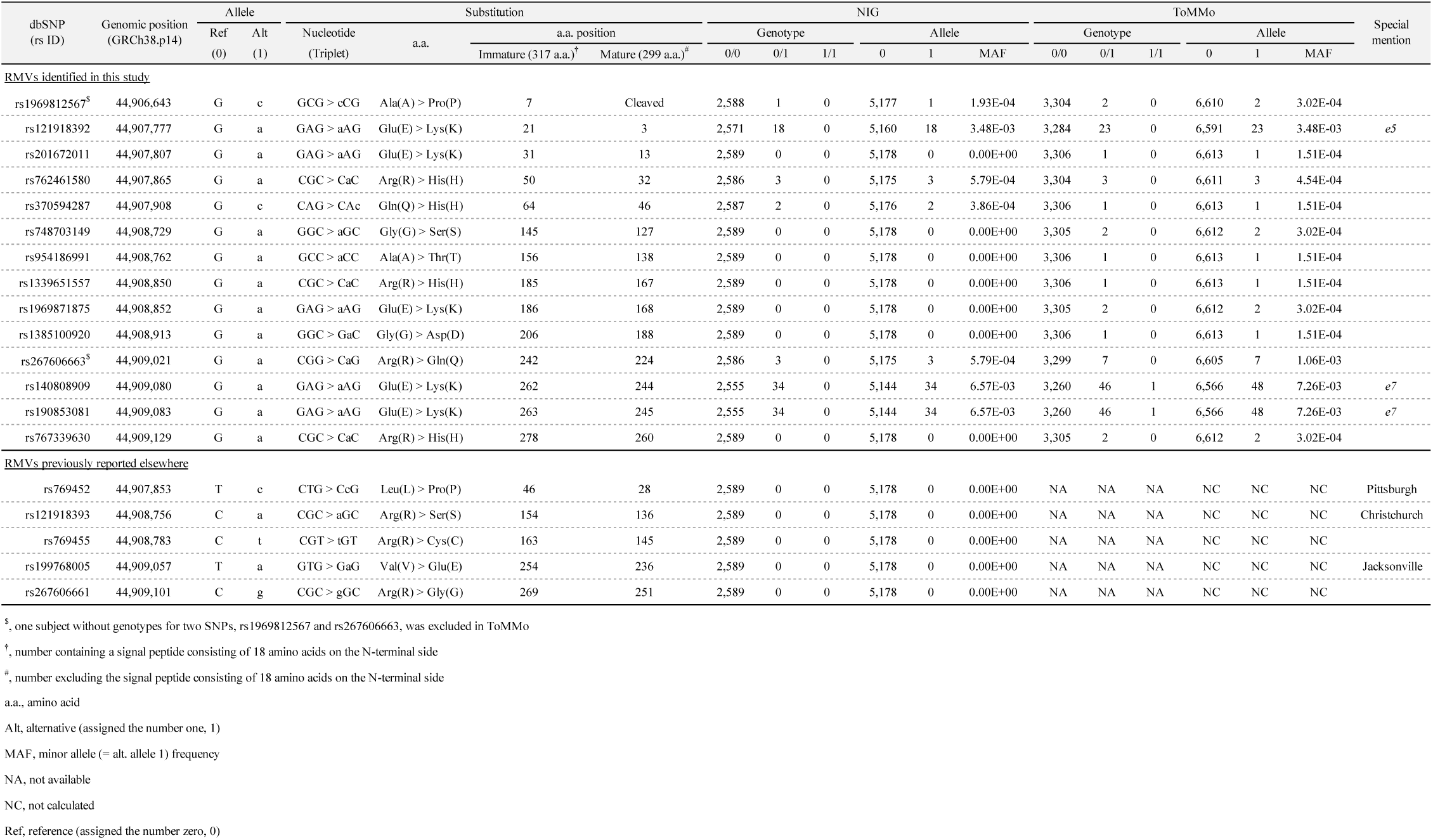
*APOE* RMVs identified.

**Table 3.**
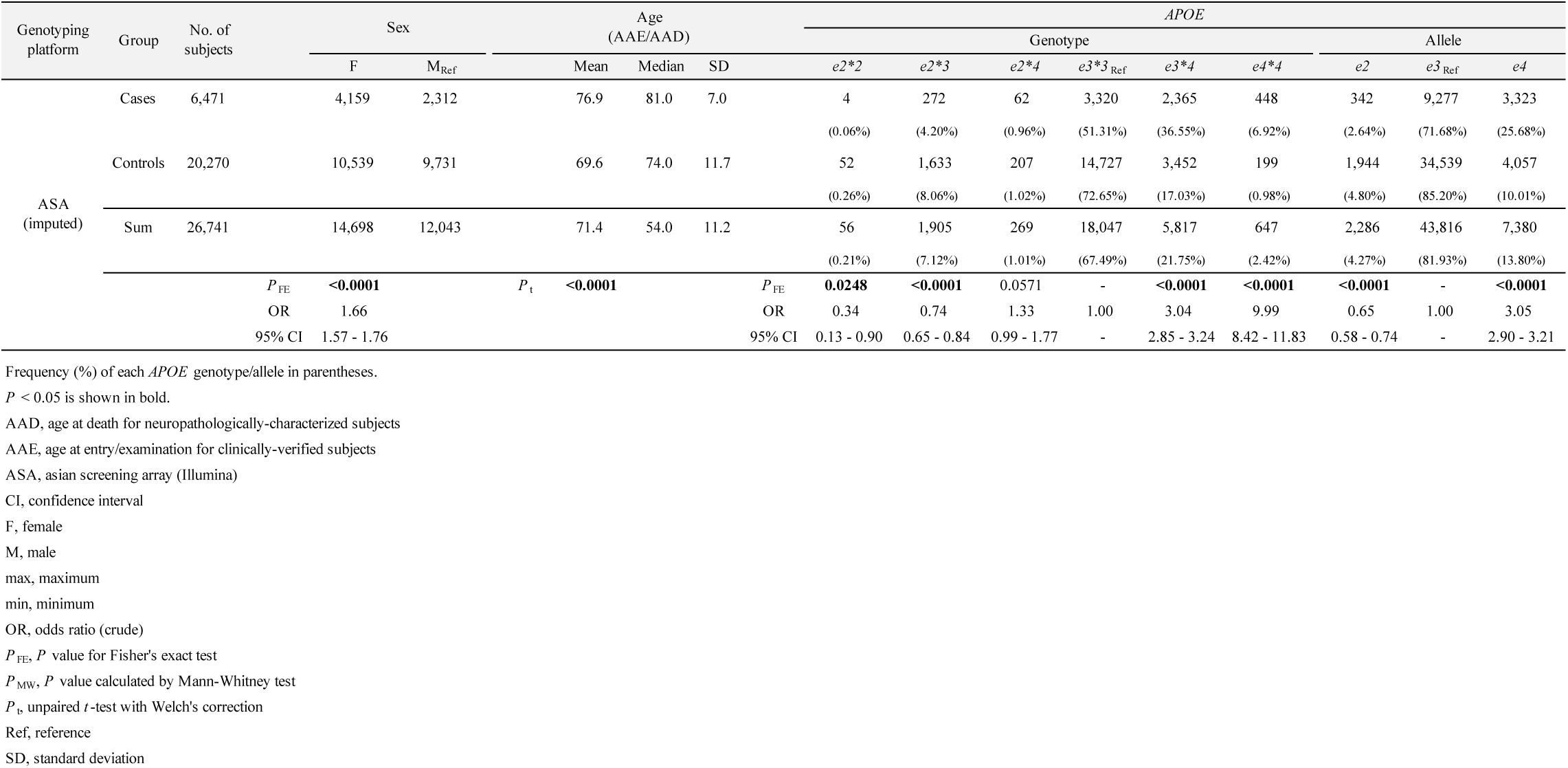
The information on subjects used in the case-control study.

None of the 14 *APOE* RMVs identified in the NIG and ToMMo cohorts were included in the ASA. Therefore, genotypes were inferred through imputation for each subject. We performed SNP imputation using Minimac4 with the Japanese reference panel, which was constructed from 1,000 Genomes Project Phase 3 data (1KGP3 [May 2013, n = 2,503]) and 3,181 Japanese whole-genome sequences from the National Center for Geriatrics and Gerontology (NCGG) biobank database. Variants with an INFO score ≥ 0.3 were included in the association analysis. Among the 14 *APOE* RMVs, genotype data were obtained for 4 variants (rs121918392 [*e5*], rs762461580, rs140808909 [*e7*], and rs190853081 [*e7*]) **(Table 4)**.

**Table 4.** Case-control study on *APOE* RMVs identified.

| dbSNP<br>(rs ID) | Allele |  | No. of subjects |  | No. of RMV carriers |  | MAF |  | *P* |  | OR | 95% CI | Special<br>mention |
| --- | --- | --- | --- | --- | --- | --- | --- | --- | --- | --- | --- | --- | --- |
|  | Ref | Alt | Cases | Controls | Cases | Controls | Cases | Controls | Unadjusted | Bonferroni<sup>†</sup> |  |  |  |
| ASA: imputed |  |  |  |  |  |  |  |  |  |  |  |  |  |
| rs121918392 | G | a | 6,413 | 19,999 | 57 | 194 | 4.41E-03 | 4.80E-03 | 5.63E-01 | 1.00E+00 | 0.92 | 0.68 - 1.23 | e5 |
| rs762461580 | G | a | 6,443 | 20,121 | 27 | 72 | 2.09E-03 | 1.81E-03 | 5.27E-01 | 1.00E+00 | 1.15 | 0.74 - 1.78 |  |
| rs140808909 | G | a | 6,402 | 19,893 | 68 | 300 | 5.26E-03 | 7.45E-03 | **8.95E-03** | **3.58E-02** | 0.70 | 0.54 - 0.92 | e7 |
| rs190853081 | G | a | 6,402 | 19,893 | 68 | 300 | 5.26E-03 | 7.45E-03 | **8.95E-03** | **3.58E-02** | 0.70 | 0.54 - 0.92 | e7 |

The OR and its 95% CI are illustrated visually in **Figure 2**.
Alt, alternative
ASA, asian screening array (Illumina)
CI, confidence interval
MAF, minor allele frequency
OR, odds ratio (adjusted)
Ref, reference
RMV, rare missense variant
<sup>†</sup>, Bonferroni-corrected *P* value, calculated by setting the number of tests to four

### TA cloning to verify RMV alleles

In one case (Subject ID #35: female, age range at death 90-99, *e3*3*) with three *APOE* RMVs (all heterozygous: rs267606663-G/a, rs140808909 [*e7*]-G/a, rs190853081 [*e7*]-G/a) **(Supplementary Table 5)**, we investigated whether these RMVs were located on the same chromosome. All the RMVs were located in exon 4 of *APOE*. Genomic DNA from the subject was used as a template for PCR, followed by TA cloning (TaKaRa Mighty TA-cloning Kit). The sequences of the primer pair used for PCR were as follows: forward primer 5 ‘-TGCCGATGACCTGCAGAA-3’, and the reverse primer was 5’-GCTGCATGTCTTCCACCAG-3’.

### Lipoprotein data collection

Measurement data for total cholesterol (TC), low-density lipoprotein cholesterol (LDL-C), high-density lipoprotein cholesterol (HDL-C), and triglycerides (TG) were available for the J-ADNI subjects included in the NIG (508 cases: baseline) and ToMMo (TC: 234 subjects; non-TC: 1,630 subjects). We examined the quantitative differences between *APOE* RMV carriers and non-carriers, if any.

### Statistical analysis

Differences in sex distribution and *APOE* genotypic and allelic distributions between cases (AD) and controls were assessed using Fisher’s exact test based on 2×2 or 2×3 contingency tables **(Table 3, Supplementary Table 4)**. Additionally, unadjusted odds ratios (OR) and 95% confidence intervals (95% CI) were calculated **(Table 3, Supplementary Table 4)**. For age, unpaired Student’s *t*-test with Welch’s correction was used **(Table 3)**.

Logistic regression analysis was conducted on *APOE* RMVs in the case-control study, with sex, age, and principal components as covariates to calculate *P*-values and ORs (95% CI) **(Table 4)**. The Bonferroni method was applied to adjust for multiple comparisons and corrections were made for the four *APOE* RMVs in the ASA analysis.

A meta-analysis was conducted to integrate and assess the association between *APOE* genotypes/alleles and AD derived from the two cohorts as described in a study by Asanomi *et al*. ^21^: the NCGG and the Japanese Genetic Study Consortium for Alzheimer’s Disease (JGSCAD) ^29^ **(Supplementary Figure 1)**. Both fixed-effects and random-effects models were assumed, and Chi-square *P*-values, pooled ORs, and 95% CIs were calculated. Cochran’s Q statistic and I² statistic were also computed to evaluate heterogeneity between the two datasets.

Comparisons of TC, LDL-C, HDL-C, TG, and the LDL-C/HDL-C (LH) ratio between the two groups (*APOE* RMV carriers vs. non-carriers) were conducted using the non-parametric Mann-Whitney *U*-test **(Supplementary Figure 4)**.

The series of genetic statistical analyses related to the case-control study **(Table 4**, **Figure 2)** were conducted using PLINK (v.1.90; https://www.cog-genomics.org/plink/). ^30^ Meta-analysis **(Supplementary Figure 1)** was conducted using StatsDirect (v.4.0.4; https://www.statsdirect.com). Other statistical analyses **(Tables 1 and 3, Supplementary Table 4, Figure 3, Supplementary Figure 4)** were performed using GraphPad Prism software (v.10.3.1; https://www.graphpad.com). *P*-values less than 0.05 were considered statistically significant.

**Figure 1.**
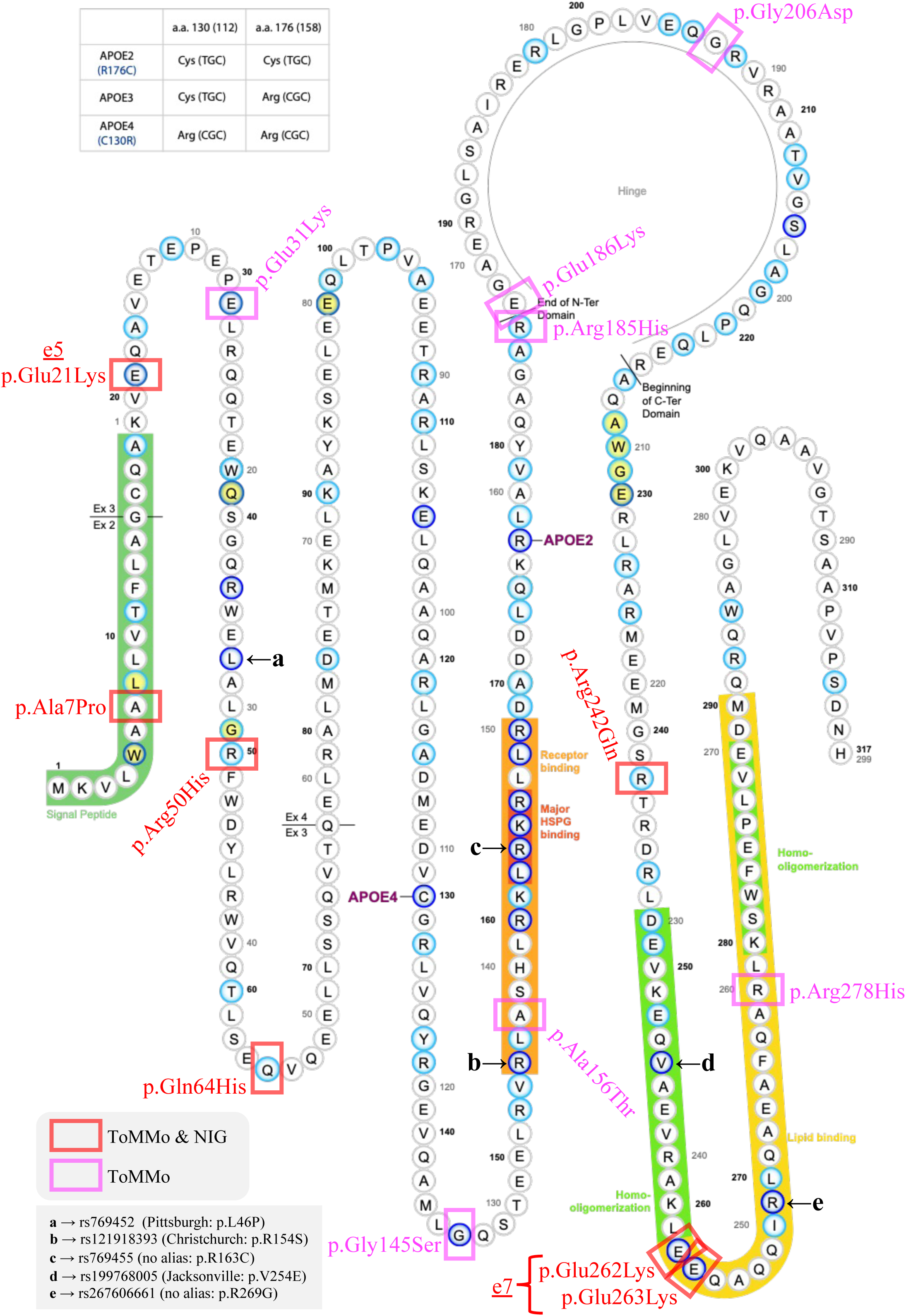
*APOE* RMVs identified in this study. The 14 *APOE* RMVs identified in this study have been added to the Alzforum MUTATIONS APOE diagram (https://www.alzforum.org/mutations/apoe). RMVs identified in both ToMMo and NIG cohorts are marked in red, whereas those identified only in ToMMo are marked in magenta.

**Figure 2.**
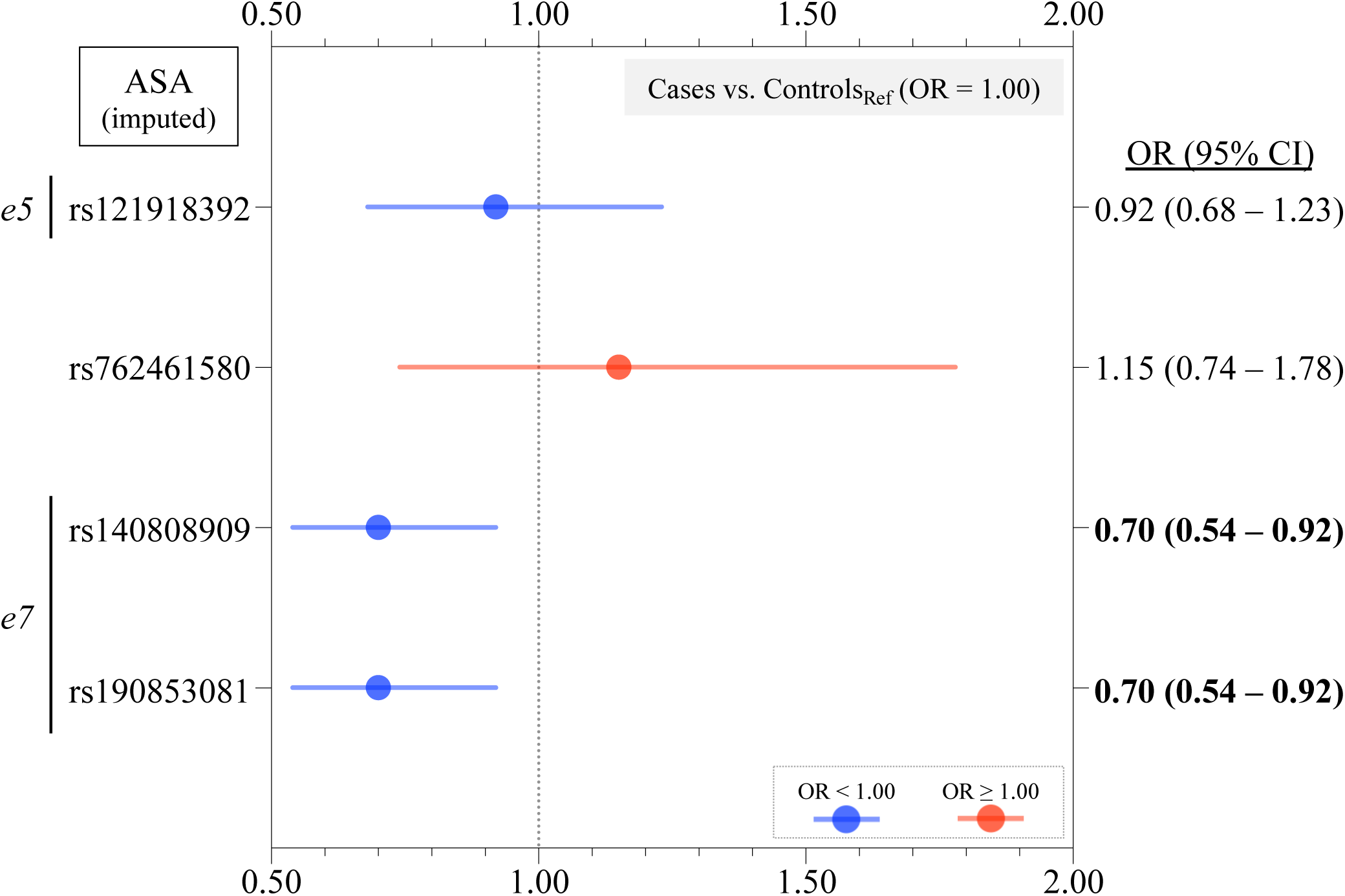
Case-control study on *APOE* RMVs identified. Of the 14 *APOE* RMVs identified in this study, four imputed variants were included in the analysis. Two RMVs, rs140808909 and rs190853081, which were in complete LD and were defined as the *e7* allele, remained statistically significant after multiple comparison corrections. The OR was < 1.0, suggesting a protective effect against AD. Detailed numerical data are presented in **Table 4**.

**Figure 3.**
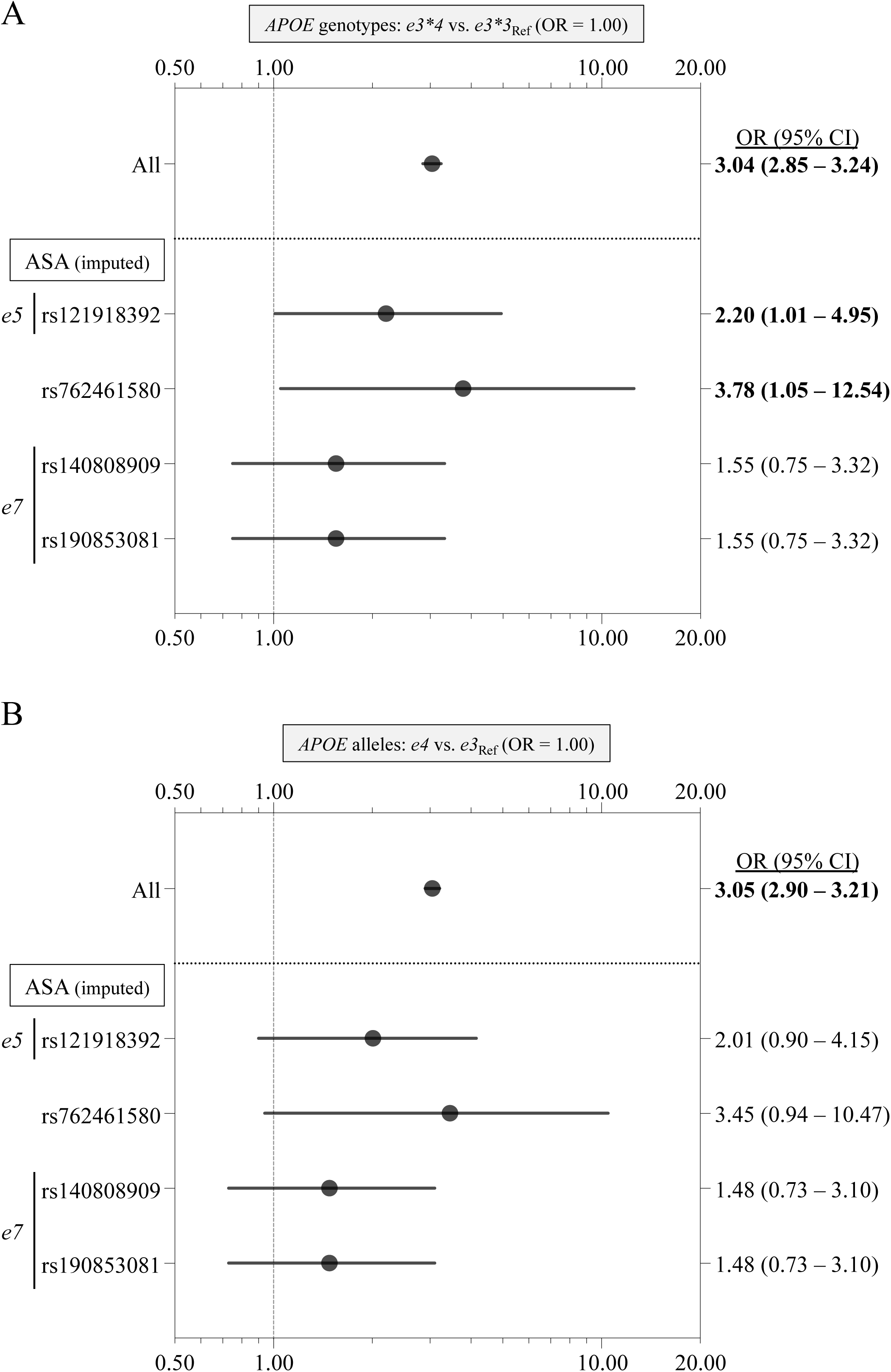
Case-control study on *APOE* genotypes and alleles in carriers with *APOE* RMVs identified. For the four *APOE* RMVs analyzed in the case-control study **(Table 4**, Figure 2), we conducted a case-control analysis focusing on subjects carrying alternative alleles, examining *APOE* genotypes **(A)** and alleles **(B)**. A forest plot showing ORs and 95% CIs is provided. For genotypes **(A)**, the OR (95% CI) of *e3*4* relative to *e3*3* was calculated. For alleles **(B)**, the OR (95% CI) of *e4* relative to *e3* was calculated. Bold text indicates that the 95% CI of the OR does not include 1.00. “All” indicates the OR and its 95% CI for all samples (6,471 AD cases and 20,270 controls) as shown in **Table 3**. Ref, reference.

### Databases

Databases and analysis websites used in this study are listed below in alphabetical order.

- Alzforum MUTATIONS APOE: https://www.alzforum.org/mutations/apoe
- ClinVar: https://www.ncbi.nlm.nih.gov/clinvar/
- dbSNP: https://www.ncbi.nlm.nih.gov/snp/
- Ensemble Human (GRCh38.p14): https://asia.ensembl.org/Homo_sapiens/Info/Index
- gnomAD (v4.1.0): https://gnomad.broadinstitute.org
- GWAS Catalog: https://www.ebi.ac.uk/gwas/home
- jMorp: https://jmorp.megabank.tohoku.ac.jp
- LDpair: https://ldlink.nih.gov/?tab=ldpair
- LDpop: https://ldlink.nih.gov/?tab=ldpop
- UCSC genome: https://genome.ucsc.edu

## Results

### Identification of APOE RMVs in the Japanese population

Using 2,589 subjects from NIG **(Table 1)**, we identified seven *APOE* RMVs **(Table 2**, **Figure 1)**: rs1969812567 (p.A7P), rs121918392 (p.E21K), rs762461580 (p.R50H), rs370594287 (p.Q64H), rs267606663 (p.R242Q), rs140808909 (p.E262K), and rs190853081 (p.E263K). All carriers were heterozygous, and no homozygotes were identified. A summary of the clinical and pathological diagnostic information of these patients is provided in **Supplementary Table 5**. Among these seven RMVs, the two variants with the highest alternative allele frequencies were rs140808909 (p.E262K) and rs190853081 (p.E263K), both with a minor allele frequency (MAF) of 0.00657 **(Table 2)**. These two variants were in complete linkage disequilibrium (LD) (LD coefficient R² = 1), indicating that they were fully co-inherited **(Supplementary Figure 2)**. Both variants were previously identified in Japanese patients with dyslipidemia and atherosclerosis, defined as the *APOE-e7* allele. ^22–24^ rs121918392 (p.E21K) was defined as the *APOE-e5* allele ^22, 25–27^ and was the third-most frequent variant after the *APOE-e7*-defining rs140808909 (p.E262K) and rs190853081 (p.E263K), with an MAF of 0.00348 **(Table 2)**. rs121918392 (p.E21K) was also associated with dyslipidemia and atherosclerosis. ^22, 25–27^

In an analysis of 3,307 subjects from the ToMMo cohort **(Table 1)**, we identified 14 *APOE* RMVs **(Table 2**, **Figure 1)**. Seven variants were identified in both the NIG and ToMMo cohorts. Additionally, seven variants were found in the ToMMo cohort but not in the NIG cohort: rs201672011 (p.E31K), rs748703149 (p.G145S), rs954186991 (p.A156T), rs1339651557 (p.R185H), rs1969871875 (p.E186K), rs1385100920 (p.G206D), and rs767339630 (p.R278H). All carriers were heterozygous, and no homozygotes were identified. For two variants in complete LD, rs140808909 (p.E262K) and rs190853081 (p.E263K), we identified one homozygous individual: sex, male; age range at entry, 60-69 years; *APOE* genotype, *e3*3*; and body mass index, 23.0. This individual had LDL-C, HDL-C, and TG levels of 96, 49, and 67, respectively, all within normal ranges (TC data were not available).

Five previously reported variants, rs769452 (p.L46P: Pittsburgh) ^13^, rs121918393 (p.R154S: Christchurch) ^14^, rs769455 (p.R163C) ^15^, rs199768005 (p.V254E: Jacksonville) ^16^, and rs267606661 (p.R269G) ^17^ **(Figure 1)**, were not detected in either NIG or ToMMo cohorts **(Table 2)**. Furthermore, these variants were not observed in the Japanese multi-omics database jMorp (54 KJPN, 38 KJPN, and 14 KJPN) ^28^ **(Supplementary Table 3)**. These variants are predominantly observed in non-East Asian populations.

### Relationship between identified APOE RMVs and common APOE alleles, e2, e3, and e4

Clarifying the common *APOE* alleles (*e2*, *e3*, and *e4*) on which each identified *APOE* RMV is located is crucial for understanding any potential genetic relationship between AD and *APOE*. Therefore, for each *APOE* RMV, we categorized subjects from the NIG and ToMMo cohorts **(Table 1)** according to the common *APOE* genotypes: *e2*2*, *e2*3*, *e2*4*, *e3*3*, *e3*4*, and *e4*4* **(Supplementary Table 2)**. The most common *APOE* genotype among *APOE* RMV carriers was *e3*3*. Several individuals were also identified with *e3*4* and *e2*3*, and one individual with *e4*4*. None of the *APOE* RMV carriers were found with *e2*2* or *e2*4*. The variant rs121918392 (p.E21K), which defines the *e5* allele, and two variants rs140808909 (p.E262K) and rs190853081 (p.E263K), which are in complete LD and define the *e7* allele, appeared to co-segregate with the *e3* allele **(Supplementary Table 2)**. The rs767339630 (p. R278H) variant was found in a subject with *e4*4*, suggesting that this variant is likely to co-segregate with the *e4* allele **(Supplementary Table 2)**.

### Ethnicity dependent frequencies of APOE RMVs

To investigate whether the 14 *APOE* RMVs identified in the NIG and ToMMo cohorts exist in other populations and to assess their frequencies, we explored public databases **(Supplementary Table 3)**. First, we examined the jMorp database. ^28^ Alternative alleles were observed in all 14 variants. For the two variants in complete LD, rs140808909 (p.E262K) and rs190853081 (p.E263K), several homozygous individuals were identified: twelve in 54 KJPN, six in 38 KJPN, and three in 14 KJPN. This result was consistent with our analysis, which showed that these two *APOE* RMVs had the highest alternative allele frequencies. The other 12 variants were observed only in heterozygous individuals.

Four variants, rs201672011 (p.E31K), rs762461580 (p.R50H), rs370594287 (p.Q64H), and rs748703149 (p.G145S), were also found in populations other than East Asian ones **(Supplementary Table 3)**. In contrast, six variants, rs121918392 (p.E21K), rs954186991 (p.A156T), rs1339651557 (p.R185H), rs267606663 (p.R242Q), rs140808909 (p.E262K), and rs190853081 (p.E263K), were highly ethnicity-dependent and observed only in the East Asian population **(Supplementary Table 3)**. Information regarding frequencies of the four variants, rs1969812567 (p.A7P), rs1969871875 (p.E186K), rs1385100920 (p.G206D), and rs767339630 (p.R278H), was not available for other populations **(Supplementary Table 3)**.

### Effect of APOE RMVs on AD

To investigate whether the 14 identified RMVs **(Table 2**, **Figure 1)** were genetically associated with AD, we conducted a case-control study involving 6,471 AD cases and 20,270 controls **(Table 3)**. Of the 14 RMVs, examined, there were four variants for which genotype data were obtained by imputation. Two variants, rs140808909 (p.E262K) and rs190853081 (p.E263K), that define the *e7* allele, showed a statistically significant association with AD **(Table 4)**. Since these two RMVs are in complete LD, they exhibited the same *P*-values and OR (95% CI): *P*_Unadjusted_ = 8.95E-03, *P*_Bonferroni_ = 3.58E-02, and OR (95% CI) = 0.70 (0.54 – 0.92) **(Table 4**, **Figure 2)**. An OR < 1.00 suggests that the *e7 allele* may have a protective effect against AD.

### Association between APOE genotypes/alleles and AD in APOE RMV carriers

By focusing on carriers of the four RMVs analyzed in the case-control study **(Table 4)**, we investigated whether *APOE* genotypes and alleles were associated with AD. Because no carriers of *APOE* RMVs with the *e2*2*, *e2*4*, or *e4*4* genotypes were identified, they were excluded from analysis. Additionally, owing to the low frequency of the *e2*3* genotype and *e2* allele, they were excluded from analysis. As a result, the comparison of genotypes focused on *e3*3* and *e3*4*, and we compared *e3* and *e4* for alleles. Results of the analyses are presented in **Figure 3** and **Supplementary Table 4**. For reference, the OR (95% CI) including both carriers and non-carriers of *APOE* RMVs, denoted as “All,” were also provided, derived from the data in **Table 3**. The OR (95% CI) for both *APOE* genotypes and alleles exhibited similar trends. Specifically, for the three RMVs defining the *e5* (rs121918392) and *e7* (rs140808909 and rs190853081) alleles, there was a tendency for reducing the risk of the *e4* allele. For example, in a comparison between the *e3*3* and *e3*4* genotypes, the OR (95% CI) for All was 3.04 (2.85-3.24), whereas it was 2.20 (1.01-4.95) for *e5* (rs121918392). In contrast, for rs762461580, there was a slight increase in risk associated with the *e4* allele. For example, in the same genotype comparison, the OR (95% CI) for All was 3.04 (2.85-3.24), but was 3.78 (1.05-12.54) for rs762461580.

### Comparison of Lipoprotein Levels Between APOE RMV Carriers and Non-Carriers

We compared blood lipoprotein levels between carriers and non-carriers of *APOE* RMVs in J-ADNI participants (508 subjects) from the NIG cohort and in participants from the ToMMo cohort (TC: 234 subjects; and non-TC: 1,630 subjects). No statistically significant differences were observed between *APOE* RMV carriers and non-carriers in terms of TC, LDL-C, HDL-C, TG, or LH ratios **(Supplementary Figure 4)**.

## Discussion

In this study, we identified 14 *APOE* RMVs among 5,896 Japanese individuals **(Table 2**, **Figure 1)**. We found that some *APOE* RMVs were specific to East Asians, including Japanese **(Supplementary Table 3)**. Notably, variants previously reported in Western populations (Christchurch ^14^ and Jacksonville variants ^16^) were completely absent in the Japanese population **(Table 2, Supplementary Table 3)**. These findings suggested that RMVs have distinct ethnic and regional specificities. To investigate their genetic association with AD, we conducted a case-control study involving 26,741 individuals **(Table 3)**. Two *APOE* RMVs, rs140808909 and rs190853081, which were in complete LD **(Supplementary Figure 2)** and defined as the *e7* allele ^22–24^, showed a statistically significant association with AD, with a protective effect **(Table 4**, **Figure 2)**. To the best of our knowledge, this association between *APOE* RMV and AD is the first of its kind in East Asians. Future studies with larger sample sizes are strongly recommended to validate these findings, as reproducibility is a critical focus. Considering the genetic diversity and ethnic specificity of the genomic variants ^31^, detailed analyses of each ethnic group are important.

In this exploratory study, we identified 14 *APOE* RMVs **(Table 2**, **Figure 1)**, with the *e7* allele ^22–24^ being the most intriguing because of its potential protective effects against AD. The two RMVs that define the *e7* allele replace two glutamate (E) residues with lysine (K) at the entry point of the lipid-binding domain on the C-terminal side of APOE **(Figure 1)**. This substitution reduces the binding affinity of e7 to LDL receptors and increases its affinity for heparin. ^32^ No previous studies have reported a genetic association between *e7* and AD. However, Youn *et al*. ^33^ suggested that the *e7* allele may be involved in cognitive impairment, based on a study of 344 amnestic patients and 345 controls in a Korean population. They identified three heterozygous carriers of the *e7* allele, all of whom belonged to the amnestic group. Two of these individuals were clinically diagnosed with AD and small vessel disease, and the remaining one was diagnosed with subjective cognitive impairment. They also predicted the three-dimensional protein structure of e7, showing that its C-terminal structure resembled that of e4, despite having an e3 background. They concluded that e7 might behave like e4 and potentially exert adverse effects on cognitive function. While our study suggests a protective effect of the *e7* allele against AD, the relationship between cognitive function and the *e*7 allele is an important area for future investigation.

We compared peripheral blood cholesterol levels (TC, LDL-C, HDL-C, TG, and LH ratios) between carriers and non-carriers of the identified *APOE* RMVs. Contrary to expectations, no significant differences were observed in cholesterol levels **(Supplementary Figure 4)**. Owing to the rarity of *APOE* RMVs, all carriers were grouped into a single category for analysis in this study. Different *APOE* RMVs may have varying effects on peripheral blood cholesterol levels; ideally, each *APOE* RMV should be analyzed separately. For instance, a study concerning the *e5* and *e7* alleles revealed a trend toward higher TC (> 220 mg/dL) and TG (> 150 mg/dL) levels compared with controls. ^34^ Considering the low population frequency of *APOE* RMVs (< 1%), comparative analyses with larger sample sizes are warranted.

This study had certain limitations. First, the number of identified RMVs **(Table 2**, **Figure 1)** was lower than expected, and further exploratory studies using a larger sample of Japanese individuals are needed. Specifically, if the focus is on late-onset AD, exploratory studies of RMVs using samples from older individuals, particularly those aged 65 and above, would be valuable. Second, we were unable to verify the reproducibility of the genetic association between AD and the *e7* allele (rs140808909 and rs190853081) in a larger independent sample of Japanese individuals, which warrants further investigation. Third, because this study was based on cross-sectional data, the longitudinal relationship between RMVs and the risk of developing AD remains unclear. Finally, functional assays of RMVs have not yet been conducted, leaving the impact of RMVs on AD pathology and the underlying molecular mechanisms unclear.

*APOE* plays a crucial role in the pathophysiology of AD, particularly in relation to Aß and tau. ^35–36^ Previous research has largely focused on the major missense alleles, *e2*, *e3* and *e4*, of *APOE*. In addition to the major *APOE* missense variants, research on *APOE* RMVs, including the Christchurch variant (rs121918393: p.R154S [p.R136S]) ^14, 37–41^, is expected to expand, further deepening our understanding of the role of APOE in AD pathophysiology. Additional studies are required to elucidate the functional significance of the 14 *APOE* RMVs identified in this study.

## Author Contributions

AM and TIkeuchi: Study design, data analysis, interpretation of data, and manuscript preparation and revision. AO, NH, TT, MH, KK, and MK: Nucleic acid extraction, quality control, sequencing, genotyping analyses, data interpretation, and manuscript revision. RM, SN, and KO: case-control studies using gene chips, data interpretation, and manuscript revision. DK, YH, TS, MTakao, RG, MTada, AK, AArakawa, MM, SM, YS, HH, TM, AAkagi, YR, HM, JS, MY, and HY provided the autopsied brains and revised the manuscript. RK and TIwatsubo: Data interpretation and manuscript revision. All authors have read and approved the final version of the manuscript.

## ACKNOWLEDGMENTS

We wish to thank all of the participants for their involvement in this study. We also gratefully acknowledge the ToMMo cohort **(Table 1)** for providing individual genomic variant data on *APOE* and its surrounding regions as well as various datasets from the subjects (research ID: 2019-0074). In the preparation of this paper, an initial rough English proofreading was performed using ChatGPT-4o, followed by proofreading by a native speaker of English.

## Supplementary Material

Supplementary figures and tables are available in the electronic version of this article.

## Data Availability

The data supporting the findings of this study are generally available upon request from the corresponding authors (AM and TIkeuchi). However, the WES data used in this study are not publicly available due to privacy or ethical restrictions.

---

**Supplementary Table 1. SS primers for all *APOE* exons and TaqMan SNP genotyping assays for rs429358 and rs7412.**

**Supplementary Table 2. *APOE* RMV carriers expanded based on six common *APOE* genotypes.**

**Supplementary Table 3. Details of 14 RMVs identified in this study and 5 RMVs previously reported elsewhere.**

**Supplementary Table 4. Case-control study on *APOE* RMVs identified.**

**Supplementary Table 5. Clinical and neuropathological diagnoses of *APOE* RMV carriers in the NIG cohort.** sample

**Supplementary Figure 1. Meta-analysis of *APOE* genotypes and alleles in NCGG and JGSCAD.**

A meta-analysis was conducted using *APOE* genotype data from two Japanese cohorts, the NCGG (Table S2) and JGSCAD (Table S4), as reported by Asanomi *et al*. ^21^ The pooled ORs and its 95% CI were calculated using both fixed effects (Fixed) and random effects (Random) models, and are presented in a forest plot. For genotype analysis **(A)**, the OR and 95% CI for five genotypes (*e2*2*, *e2*3*, *e2*4*, *e3*4*, *e4*4*) were calculated in comparison to *e3*3*. For allele analysis **(B)**, the OR and 95% CI for two alleles (*e2* and *e4*) were calculated in comparison to *e3*. The OR and 95% CI are highlighted in bold if the 95% CI does not include 1.00. Ref, reference.

**Supplementary Figure 2. Pairwise LD measures between two *APOE* RMVs, rs140808909 and rs190853081, determining the *e7* allele.**

Using the LDpair Tool on the LDlink website (https://ldlink.nih.gov/?tab=ldpair), we analyzed the haplotypes and their frequencies, as well as the strength of LD (D’ and R²) between the two RMVs, rs140808909 and rs190853081, in the East Asian (EAS) population (504 subjects). Sub-analyses were conducted separately for the Japanese in Tokyo, Japan (JPT, 104 subjects) and the Southern Han Chinese (CHS, 105 subjects) populations. Three A-A haplotypes were observed in the EAS population, one in JPT, and two in CHS. Notably, these two RMVs were not detected in populations outside the EAS database.

**Supplementary Figure 3. Sequence verification in an individual carrying three *APOE* RMVs: p.Arg242Gln (rs267606663-G/a), p.Glu262Lys (rs140808909-G/a), and p.Glu263Lys (rs190853081-G/a).**

Subject ID #35 (*APOE* genotype, *e3*3*; **Supplementary Table 5**) harbored three RMVs. The results of SS of the genomic DNA are shown in **A**. Three RMVs (rs267606663, rs140808909, and rs190853081) were detected, all of which were G/A heterozygotes. PCR products generated using genomic DNA as a template were cloned into a TA vector and SS of the plasmid DNA was performed. The results are presented in Section **B**. It was found that the alternative allele “a” of rs267606663 (G/a) does not co-segregate with the alternative alleles “a” of the other two RMVs, rs140808909 (G/a) and rs190853081 (G/a), and is located independently on a different chromosome. Asterisks indicate nucleotide substitutions; black represents the reference allele, and red represents the alternative allele.

**Supplementary Figure 4. A comparison of TC, LDL-C, HDL-C, and TG levels and L/H ratios between *APOE* RMV carriers and non-carriers in the J-ADNI and ToMMo cohorts.**

Each measurement item is represented as a box-and-whisker plot (whiskers, 5th to 95^th^percentiles; +, mean) divided into J-ADNI and ToMMo groups and further subdivided into *APOE* RMV carriers and non-carriers. Numbers in parentheses indicate the sample size for each group. The light blue vertical lines indicate reference values for each measurement item: **A**, TC 150–219 mg/dL; **B**, LDL-C 70–139 mg/dL; **C**, HDL-C 40–96 mg/dL [females 40–96 mg/dL, and males 40–86 mg/dL]; **D**, TG 50–149 mg/dL; and **E**, LH ratio ≤1.5. No significant differences were observed in any of these comparisons. *P*_MW_, *P*-value of Mann-Whitney *U*-test.

