## Supplementary figures and images for "Association of rare *APOE* missense variants with Alzheimer’s disease in the Japanese population"

### Supplemental Figure 1

Supplementary Fig. 1. Meta-analysis of *APOE* genotypes and alleles in NCGG and JGSCAD

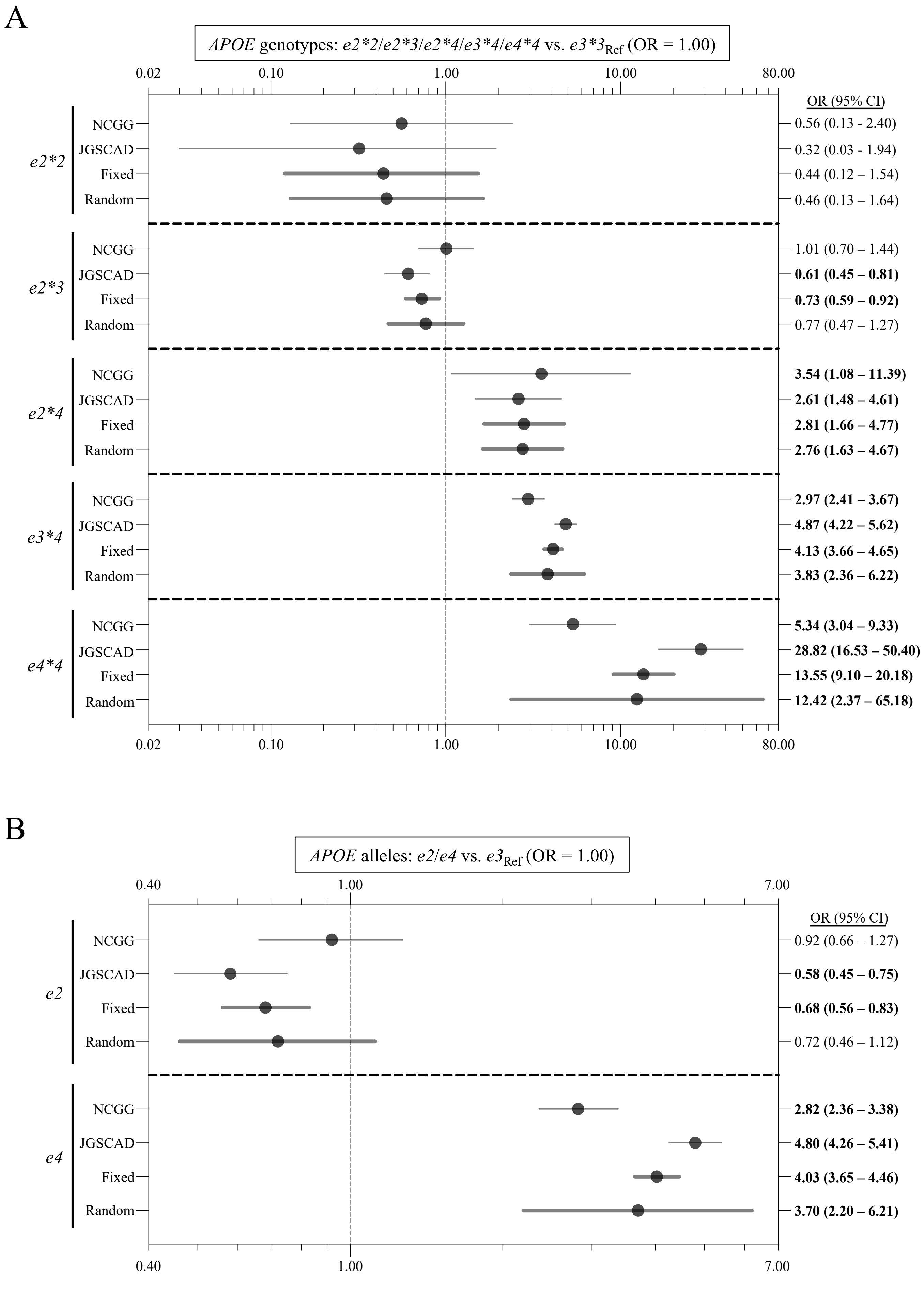
