## Supplemental Figure 2 for "Association of rare *APOE* missense variants with Alzheimer’s disease in the Japanese population"

Supplementary Fig. 2. Pairwise LD measures between two *APOE* RMVs, rs140808909 and rs190853081, determining the *e7* allele.

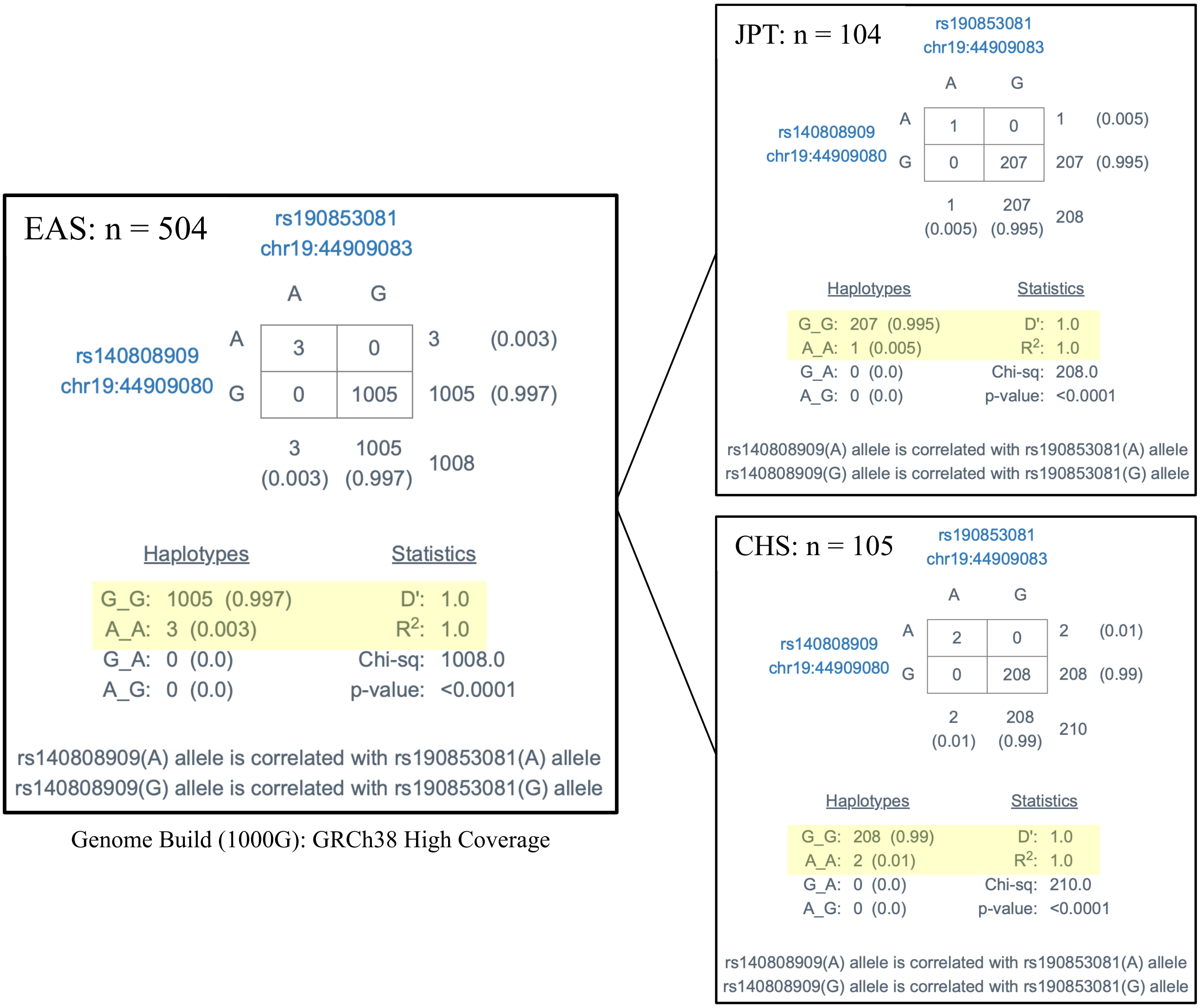
