## Supplemental Figure 3 for "Association of rare *APOE* missense variants with Alzheimer’s disease in the Japanese population"

Supplementary Fig. 3. Sequence verification on an individual who carries the *APOE* RMV, exhibiting heterozygosity for both the p.Arg242Gln and p.Glu262Lys-p.Glu263Lys variants

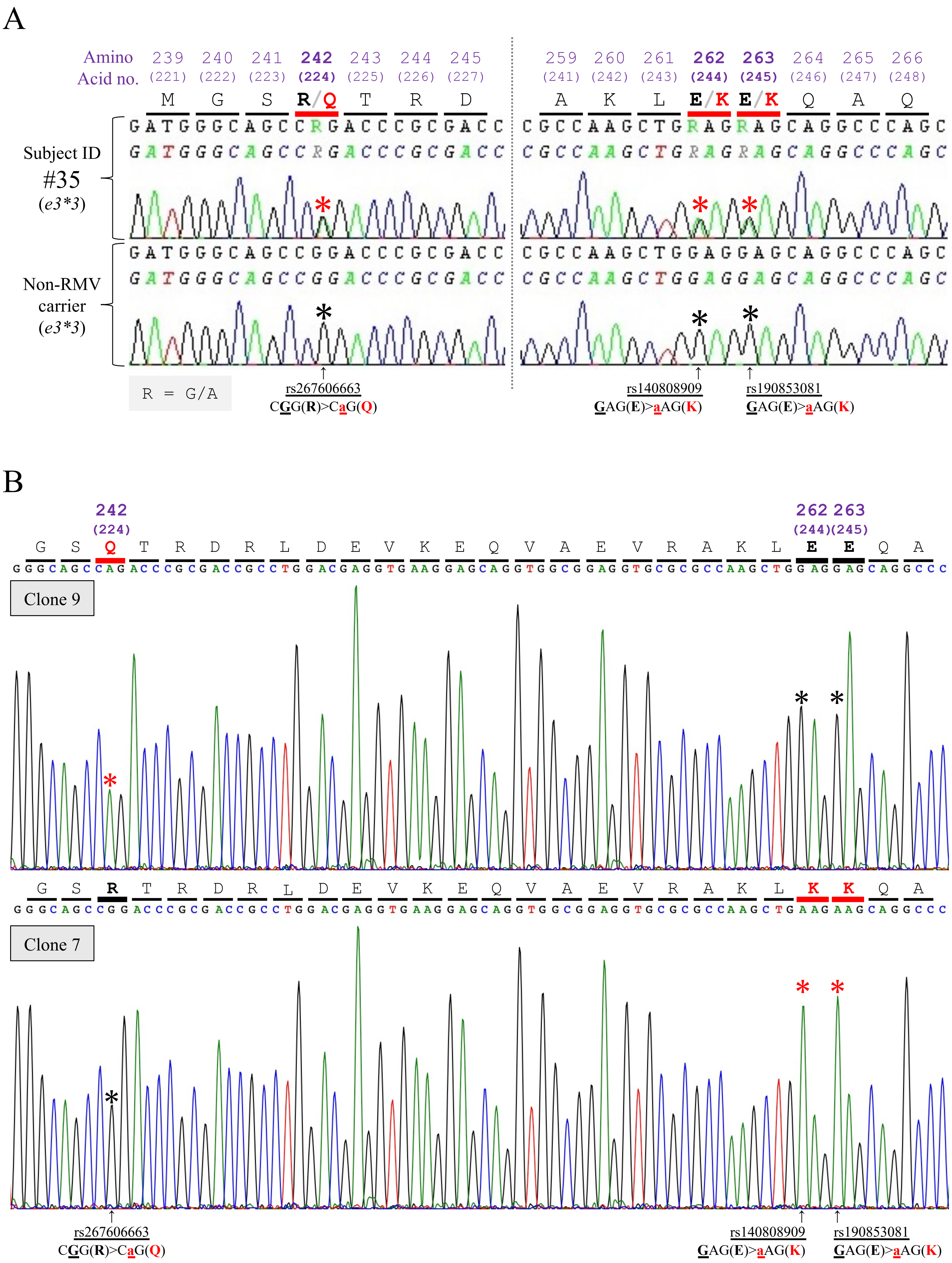
