## Supplemental Figure 4 for "Association of rare *APOE* missense variants with Alzheimer’s disease in the Japanese population"

Supplementary Fig. 4. A comparison of TC, LDL-C, HDL-C, and TG levels between *APOE* RMV carriers and non-carriers in the J-ADNI and ToMMo cohorts

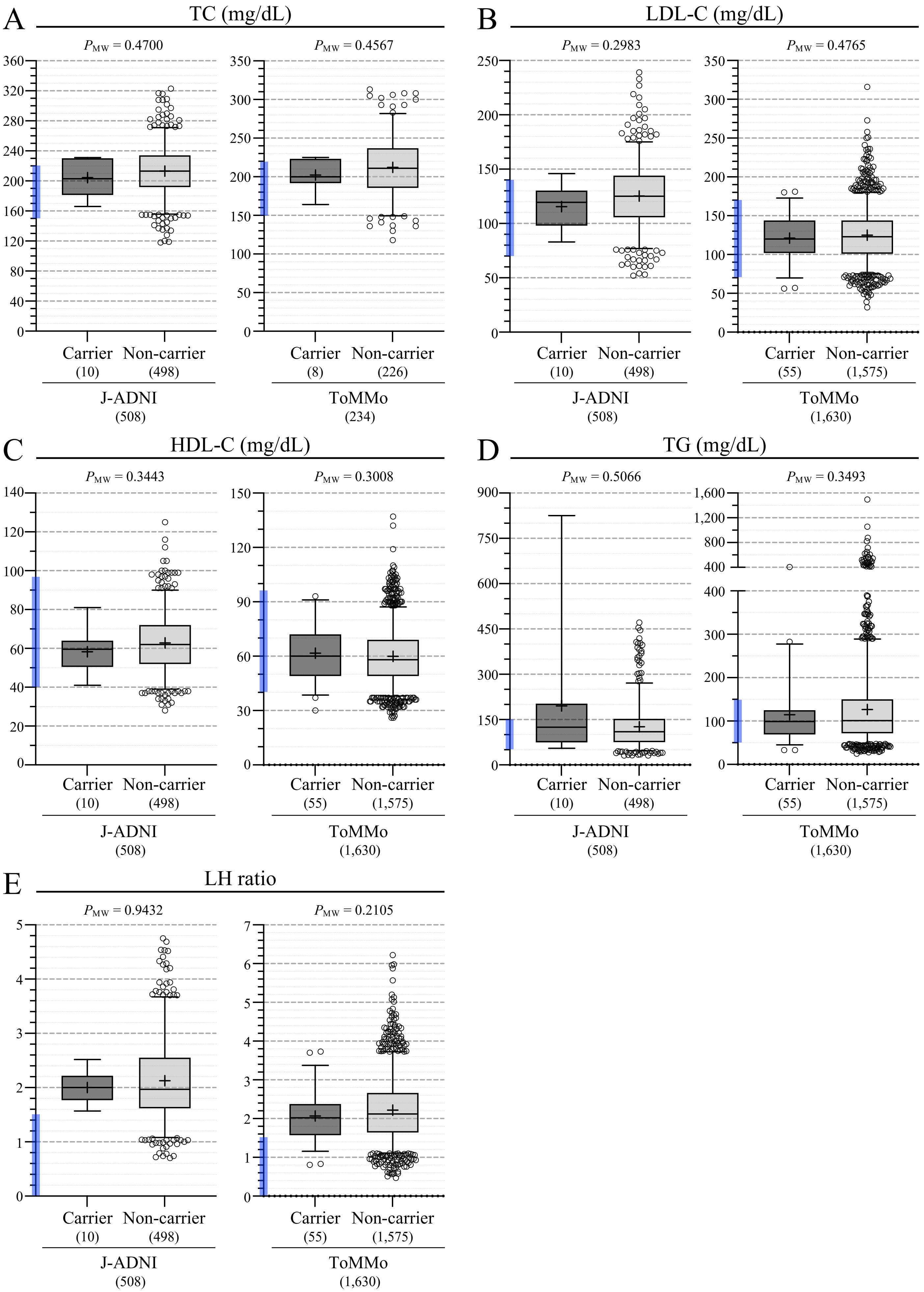
